# Get With The Guidelines-Heart Failure Hospital Participation and its Association with Guideline-Directed Medical Therapy and Outcomes

**DOI:** 10.64898/2026.03.03.26347559

**Authors:** Aradhana Verma, Gregg C Fonarow, Paul Heidenreich, Larry A. Allen, Andrew P. Ambrosy, Shun Kohsaka, Anubodh S. Varshney, Nicholas Brownwell, Jun Fan, Alexander T. Sandhu

## Abstract

**Purpose:** Despite strong evidence, real-world adoption of guideline-directed medical therapy (GDMT) for heart failure with reduced ejection fraction (HFrEF) remains suboptimal. The Get With The Guidelines–Heart Failure (GWTG-HF) program was designed to close gaps in care. We evaluated whether hospital participation in GWTG-HF is associated with greater GDMT intensity and improved outcomes.

**Methods:** We conducted a retrospective analysis (2013-2021) of Medicare beneficiaries with Part A and Part D hospitalized with HFrEF. Using a multiple baseline time series design, we compared changes in GDMT prescribing and outcomes at hospitals before and after GWTG-HF enrollment with hospitals that never participated. The primary outcome was a 90-day post-discharge prescription-fill GDMT score summarizing use and dose of beta blockers, renin-angiotensin system inhibitors (RASI; ACE inhibitor/ARB/ARNI), and mineralocorticoid receptor antagonists (MRA). Secondary outcomes included class-specific medication fills, achievement of ≥50% target doses, and 30-day, 90-day, and 1-year all-cause and HF readmission and mortality. We adjusted for baseline hospital performance, patient characteristics, and temporal trends.

**Results:** Among 1,274,863 Medicare beneficiaries hospitalized for HFrEF, 53.5% were treated at hospitals that never participated in GWTG-HF and 9.6% at hospitals that joined GWTG-HF before hospitalization. Unadjusted median GDMT scores increased from 3.0 in both groups to 4.0 in non-participating hospitals and 4.5 in GWTG-HF hospitals at 90 days (p<0.001). Hospital enrollment was associated with a higher 90-day GDMT score (+0.15 points; 95% CI 0.12-0.18; p<0.001), and greater use of beta blockers, RASI, and MRA, but not ARNI. HF readmission did not differ significantly; however, GWTG-HF participation was associated with lower all-cause mortality at 30 days (OR 0.95; 95% CI:0.92-0.98), 90 days (OR: 0.97; 95% CI: 0.95-0.99), and 1 year (0.97; 95% CI: 0.95-.0.99; all p<0.05).

**Conclusion:** Hospital participation in GWTG-HF was associated with higher GDMT intensity and lower mortality, supporting structured quality programs to improve HFrEF care.

## Introduction

The Get With The Guidelines-Heart Failure (GWTG-HF) registry is a national, hospital-based quality improvement initiative launched in 2005 to enhance heart failure (HF) care by promoting adherence to accepted treatment guidelines.^1,2^ The program includes workshops, webinars, hospital tool kits, performance benchmarks, and achievement awards to drive high-quality HF care.^3^ While prior analyses demonstrated GWTG-HF hospitals have higher performance on process of care measures compared to non-participating hospitals, the association between joining the GWTG-HF registry and subsequent within-hospital changes in HF quality of care and outcomes have not been previously evaluated.^4^

The primary objective of this study was to evaluate the association between hospital enrollment in GWTG-HF and quality of care for patients with HF with reduced ejection fraction (HFrEF) based on guideline-directed medical therapy (GDMT) utilization and clinical outcomes. Using national Medicare claims data linked to hospital GWTG-HF participation data, we compared changes in HF quality of care and outcomes over time with a multiple baseline time series analysis between hospitals that joined the GWTG-HF registry and those that never participated between 2013 and 2021.

## Methods

### Study Design

We used a multiple baseline time series design to assess the association between GWTG-HF participation and outcomes (Figure 1). This quasi-experimental approach is conceptually similar to a controlled interrupted time series analysis, but is well-suited for interventions introduced in a staggered fashion across different sites at different time points as compared with a single intervention implemented simultaneously across sites.^5–6^ In this design, each hospital serves as its own comparator prior to enrollment, while hospitals not yet enrolled and those that never participated in GWTG-HF during the study period served as concurrent controls, allowing separation of the intervention from underlying secular trends. The staggered enrollment of hospitals into GWTG-HF at varying time points strengthens the analysis by providing multiple “interruption” points and reducing bias from time-varying effects common to all sites. This approach has been previously used to evaluate system-level healthcare interventions introduced sequentially across institutions and provides a robust framework for assessing real-world quality improvement initiatives.^5,7^

**Figure 1:**
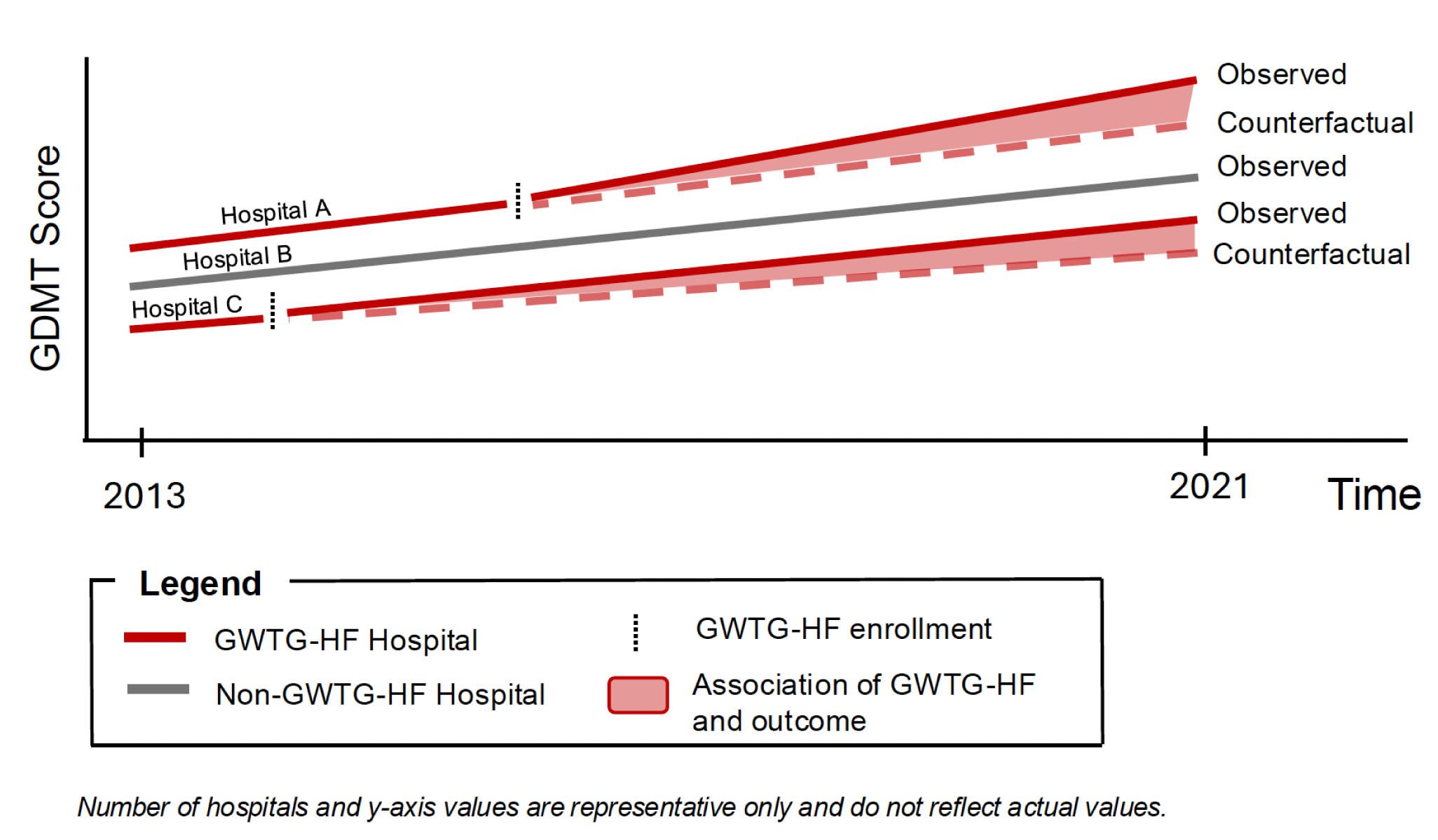
Schematic of the multiple baseline time series design showing staggered GWTG-HF enrollment across hospitals compared with the non-GWTG-HF control arm.

### Study Population

We analyzed data from Medicare fee-for-service claims from January 1, 2011 to December 31, 2022. We identified hospitalizations for HFrEF between 2013 and 2021 based on a principal discharge diagnosis of HF in a short-term acute care hospital. Patients were classified as having HFrEF if they had only systolic heart failure ICD codes during the index hospitalization.^8^ Among participants with systolic and diastolic HF codes during their hospitalization, patients were classified as HFrEF if the majority of HF codes documented in a one-year lookback period were systolic HF codes.

We restricted the analysis to individuals with continuous enrollment in Medicare Parts A for at least 1 year to ensure capture of baseline comorbidities. We also required enrollment in Medicare Part D (outpatient prescription plan) for at least 6 months prior to hospitalization to capture pre-admission GDMT and for at least 6 months post-discharge or until death.

We excluded patients with a history of heart transplant, left ventricular assist device implantation, advanced chronic kidney disease (CKD) stage 4 or 5, or dialysis dependence given these clinical characteristics impact the benefits of GDMT. We also excluded individuals who were transferred to another acute-care short-stay hospital, died during hospitalization, or were discharged to hospice. We included all eligible hospitalizations in the analysis, allowing multiple hospitalizations for an individual Medicare patient.

### Exposure

The primary exposure was hospitalization at a facility with active GWTG-HF registry participation. Patients were classified based on the participation status of the admitting hospital at the time of hospitalization. Specifically, patients admitted during periods when their hospital was actively enrolled in the GWTG-HF registry were classified as GWTG-HF patients, whereas those admitted to hospitals during periods without active registry participation were classified as non-GWTG-HF. Because hospitals enrolled in GWTG-HF at different times throughout the study period, the number of participating hospitals increased over time (Figure S1). Individual hospitals could contribute observations to both exposure groups, contributing patients to the non-GWTG-HF group prior to enrollment and to the GWTG-HF group after enrollment.

We excluded patients treated at hospitals that enrolled in GWTG-HF prior to the study period since we lacked data on hospital quality of care prior to their enrollment in the GWTG-HF registry. We also excluded patients at hospitals that had previously participated in GWTG-HF but disenrolled prior to the hospitalization.

### Outcomes

The primary outcome was a composite GDMT score at 90 days post-discharge reflecting both use and dose intensity of evidence-based therapies for HFrEF, adapted from the continuous 4-Pillar Score.^9^ Beta blockers, angiotensin-converting enzyme inhibitor (ACEi)/angiotensin II receptor blockers (ARB), and mineralocorticoid receptor antagonists (MRA) contributed 2 points for therapy initiation plus 2 points multiplied by the proportion of target daily dose achieved, while angiotensin receptor-neprilysin inhibitors (ARNI) contributed 4 points for initiation plus 2 points multiplied by the target daily dose. This score reflects the goals of both initiation of therapy and dose uptitration, which are noted in the ACC/AHA Clinical Performance Measures for HF.^10^ We have also previously demonstrated the association between this score and subsequent clinical outcomes.^9^ We did not include sodium glucose cotransporter 2 inhibitors (SGLT2i) in the primary outcome given the majority of the study period preceded their approval for HF. The total score was a sum of these components, ranging from 0 to 14 (Figure S2).

For each drug class, we identified the last prescription fill within 90 days post-discharge. We used the 90-day window post-discharge because shorter time windows may fail to capture medications in which the patient still has the remainder of an active medication filled prior to admission. This approach does potentially incorporate post-discharge GDMT optimization in addition to inpatient GDMT optimization; however, prior studies have illustrated very low rates of post-discharge medication optimization.^11^ We also evaluated the GDMT score at 180 days post-discharge. For patients who died or were rehospitalized during the time window, we still used the last medication fill.

Secondary outcomes included the composite GDMT score for quadruple therapy including SGLT2i; binary indicators for the use of each GDMT medication class; and binary indicators for use of ≥50% of target dose for beta blockers and renin angiotensin aldosterone system inhibitors (RASI).^10^ Finally, we measured clinical outcomes of HF readmission, all-cause readmission, and death at 30 days, 90 days, and one year post-discharge. HF readmission was identified as an acute care hospitalization with a principal diagnosis of HF.

### Study Variables

Patient-level predictor variables included sociodemographic and clinical characteristics. Sociodemographic variables included age, sex, race/ethnicity, rural/urban classification, and CDC Social Vulnerability Index (SVI) based on zip code. We identified medical comorbidities using a one-year lookback period prior to hospitalization based on ICD-9^th^ and ICD-10^th^ revision diagnostic codes. Comorbidities included cardiovascular disease (prior HF, myocardial infarction, cerebrovascular disease/transient ischemic attack, peripheral arterial disease) and non-cardiac disease (cancer, chronic kidney disease, chronic obstructive pulmonary disease, diabetes mellitus, liver disease, frailty). Hospital-level characteristics included geographic region, bed size, teaching status, and CMS quality (range 1-5).

### Statistical Analysis

We compared sociodemographic, hospitalization characteristics, comorbidities, and pre-admission HF medications for patients stratified by hospital GWTG-HF participation. In addition, we compared hospital characteristics between hospitals that participated in GWTG-HF and those that never participated during the study period. Given the large sample size, we primarily evaluated the magnitude of differences between groups using standardized mean differences (SMD). SMD of 0.2, 0.5, 0.8 were considered small, medium, and large differences, respectively.^12^ We also compared groups with Chi-squared tests for categorical variables and t-tests for continuous variables.

We used a mixed effects model with a random intercept for the discharging hospital to account for baseline outcome levels for each hospital and a continuous measure of each quarter-year in the study period to capture secular trends. For the primary analysis, we evaluated a level change (abrupt change in the temporal trend) in the outcome after a hospital joined the GWTG-HF registry. We also evaluated the slope change (change over time after joining GWTG-HF) as a secondary analysis. We tested whether there were significant differences between temporal trends in GDMT prescription for non-GWTG-HF hospitals and GWTG-HF hospitals before enrolling in GWTG-HF. Because we found no significant difference, we assumed parallel pre-intervention trends for the primary analysis (Table S1). As a sensitivity analysis, we repeated the primary analysis while modeling different baseline slopes for GWTG-HF and non-GWTG-HF hospitals prior to registry participation.

We adjusted all models for patient age, sex, race/ethnicity, rurality, CDC SVI quartile, comorbidities, and seasonal variation. For the GDMT score, a continuous outcome, we used mixed effects linear regression. For individual medications, readmission, and mortality, we used mixed effects logistic regression.

We performed multiple sensitivity analyses. First, we analyzed differences in the GDMT score and clinical outcomes without adjustment for patient characteristics and without the hospital-level random intercept. Second, we repeated the analysis without the hospital-level intercept with adjustment for hospital characteristics. Third, we repeated the primary analysis while excluding those who died before the 90-day follow-up. We applied a p-value threshold of <0.05 as statistically significant.

## Results

Among 1,274,863 Medicare beneficiaries with Parts A and D hospitalized for HFrEF, 53.5% (n= 681,935) were treated at hospitals that never participated in GWTG-HF and 9.6% (n=122,554) at hospitals that joined GWTG-HF during the study period (Figure 2). The remaining patients were excluded because their hospital had withdrawn from the GWTG-HF registry prior to the hospitalization (17.6%, n=224,025) or their hospital joined the GWTG-HF registry before the study period (19.3%, n=246,349).

**Figure 2:**
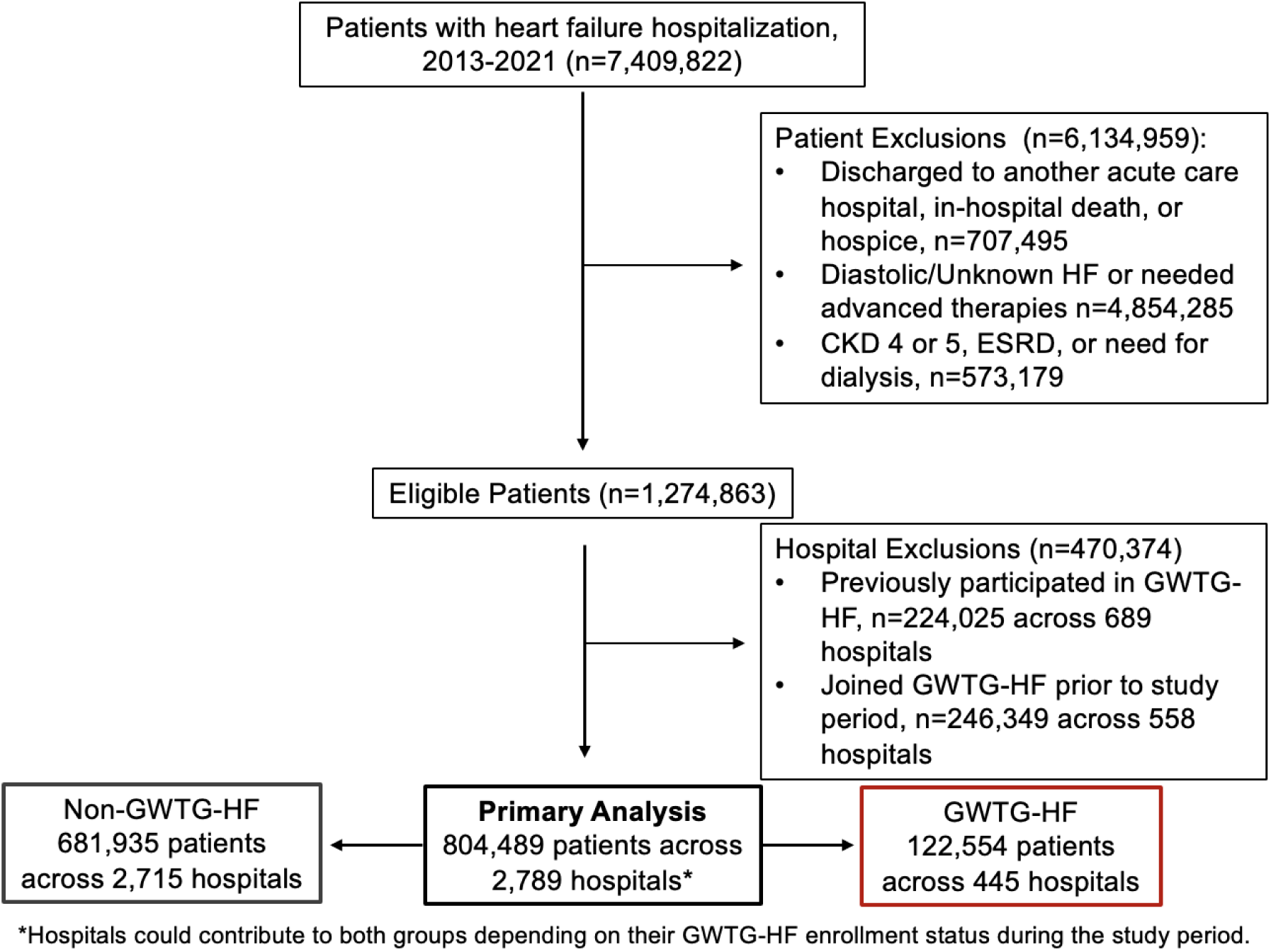
Inclusion and exclusion criteria for primary analysis.

Patient sociodemographic and clinical data are presented in Table 1. The average age was 77 years and about 42.8% were female. The majority of patients were White (69.0%), followed by Black or African American (19.6%), Hispanic or Latino (8.4%), Asian or Pacific Islander (1.6%), and American Indian or Alaskan Native (0.5%). Nearly one in five patients resided in rural areas, 21.6% of patients in Non-GWTG-HF hospitals compared with 10.2% in GWTG-HF hospitals (p<0.001, SMD 0.3). Patients hospitalized in non-GWTG hospitals were more likely to reside in communities with moderate-high social vulnerability (CDC SVI 0.51-0.75: 27.9% vs. 23.0%) compared with GWTG-HF hospitals; the proportion at the highest social vulnerability was similar (CDC SVI 0.76-1.00 31.8% vs. 30.9%). Cardiac comorbidities were common in both groups, including hypertension (96.5%), coronary artery disease (79.5%), and atrial fibrillation/flutter (62.4%). Non-cardiovascular disease was also similar across groups and highly prevalent, such as diabetes (56.6%), chronic kidney disease (56.4%), frailty (21.1%), and liver disease (19.9%).

**Table 1:**
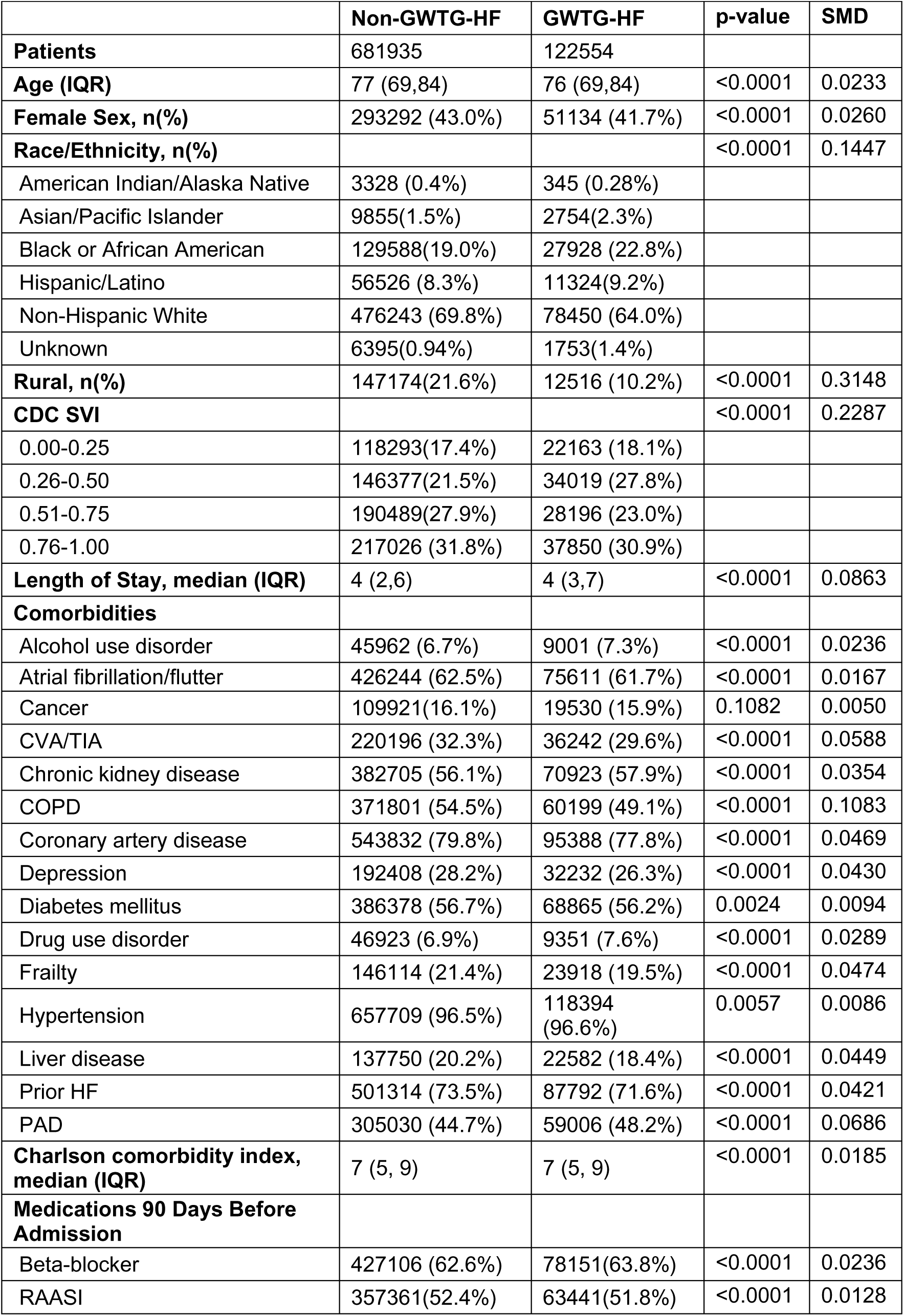
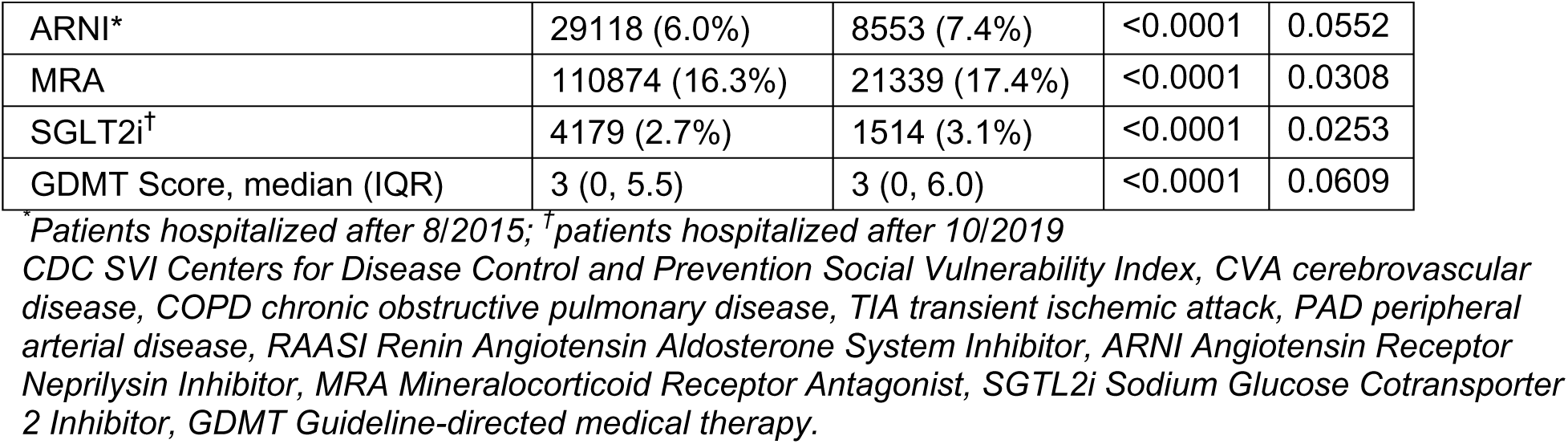
Sociodemographic data, clinical characteristics, and pre-admission HF therapies stratified by patients hospitalized in GWTG-HF versus Non-GWTG-HF participating hospitals.

Baseline HF therapies in the 90 days prior to hospitalization were similar across both groups with an unadjusted median GDMT score of 3.0 [IQR: 0-5.5] (SMD 0.1). Rates of beta blocker (63.8% vs. 62.6%), RASI (51.8% vs. 52.4%), and MRA (17.4 vs. 16.3%) were also similar between GWTG-HF and non-GWTG-HF hospitals (SMDs of 0.02, 0.01, and 0.03, respectively).

Hospital characteristics are presented in Table 2. There were 445 hospitals that had joined the GWTG-HF registry during the study period and 2,344 hospitals that did not join the registry during the study period. Supplement Figure 1 displays the overall pattern of hospitals joining the GWTG-HF registry over time. Non-GWTG-HF hospitals were more likely to be small hospitals (<100 beds; 42.3% vs. 13.3%) whereas GWTG-HF hospitals were more likely to be large (>400 beds; 25.4% vs. 8.2%). GWTG-HF hospitals were more likely to be teaching hospitals (74.4% vs. 37.3%, SMD 0.87).

**Table 2:** Hospitals characteristics stratified by GWTG-HF participation during study period.

|  | **Non-GWTG-HF** | **GWTG-HF** | **p-value** | **SMD** |
| --- | --- | --- | --- | --- |
| **No of hospitals, n** | 2344 | 445 |  |  |
| **Number of beds, n(%)** |  |  | <0.0001 | 0.9286 |
| <100 | 992 (42.3%) | 59 (13.3%) |  |  |
| 100-400 | 967 (41.3%) | 272 (61.1%) |  |  |
| >400 | 191 (8.2%) | 113(25.4%) |  |  |
| Missing | 194 (8.3%) | 1 (0.22%) |  |  |
| **Region, n(%)** |  |  | <0.0001 | 0.5511 |
| Northeast | 300 (12.8%) | 91(20.5%) |  |  |
| Midwest | 508 (21.7%) | 112 (25.2%) |  |  |
| South | 972 (41.5%) | 127 (28.5%) |  |  |
| West | 370 (15.8%) | 114 (25.6%) |  |  |
| Missing | 194 (8.3%) | 1 (0.22%) |  |  |
| **Teaching Hospital, n(%)** |  |  | <0.0001 | 0.7660 |
| Yes | 959 (37.3%) | 331 (74.4%) |  |  |
| No | 1191 (50.8%) | 113 (25.4%) |  |  |
| Missing | 194 (8.3%) | 1 (0.22%) |  |  |
| **CMS quality rating** |  |  | <0.0001 | 0.7738 |
| 1 | 136 (5.8%) | 31 (7.0%) |  |  |
| 2 | 369 (15.7%) | 95 (21.4%) |  |  |
| 3 | 525 (22.4%) | 133 (29.9%) |  |  |
| 4 | 447 (19.1%) | 99 (22.3%) |  |  |
| 5 | 223 (9.5%) | 77 (17.3%) |  |  |
| Missing | 644 (27.5%) | 10 (2.3%) |  |  |

*Hospitals that participated exclusively in GWTG-HF or as a non-GWTG-HF hospital during the study period were included.*

### Guideline-Directed Medical Therapy Fills After Discharge

Based on Medicare Part D prescription fills at 90 days following hospitalization, the median GDMT score increased to 4.0 [IQR 2.3-6.5] in non-GWTG-HF hospitals and 4.5 [IQR 2.3-7.0] in GWTG-HF hospitals at 90 days post-discharge (p<0.001). At 180 days post-discharge, the median GDMT score increased to 4.8 [IQR 2.3-7.0] in non-GWTG hospitals and 5.1 [IQR 2.5-7.8] in GWTG-HF hospitals (p<0.001) (Table 3). Figure 3 displays the average post-discharge GDMT score across hospitals per month, relative to hospital enrollment in the GWTG-HF registry.

**Figure 3:**
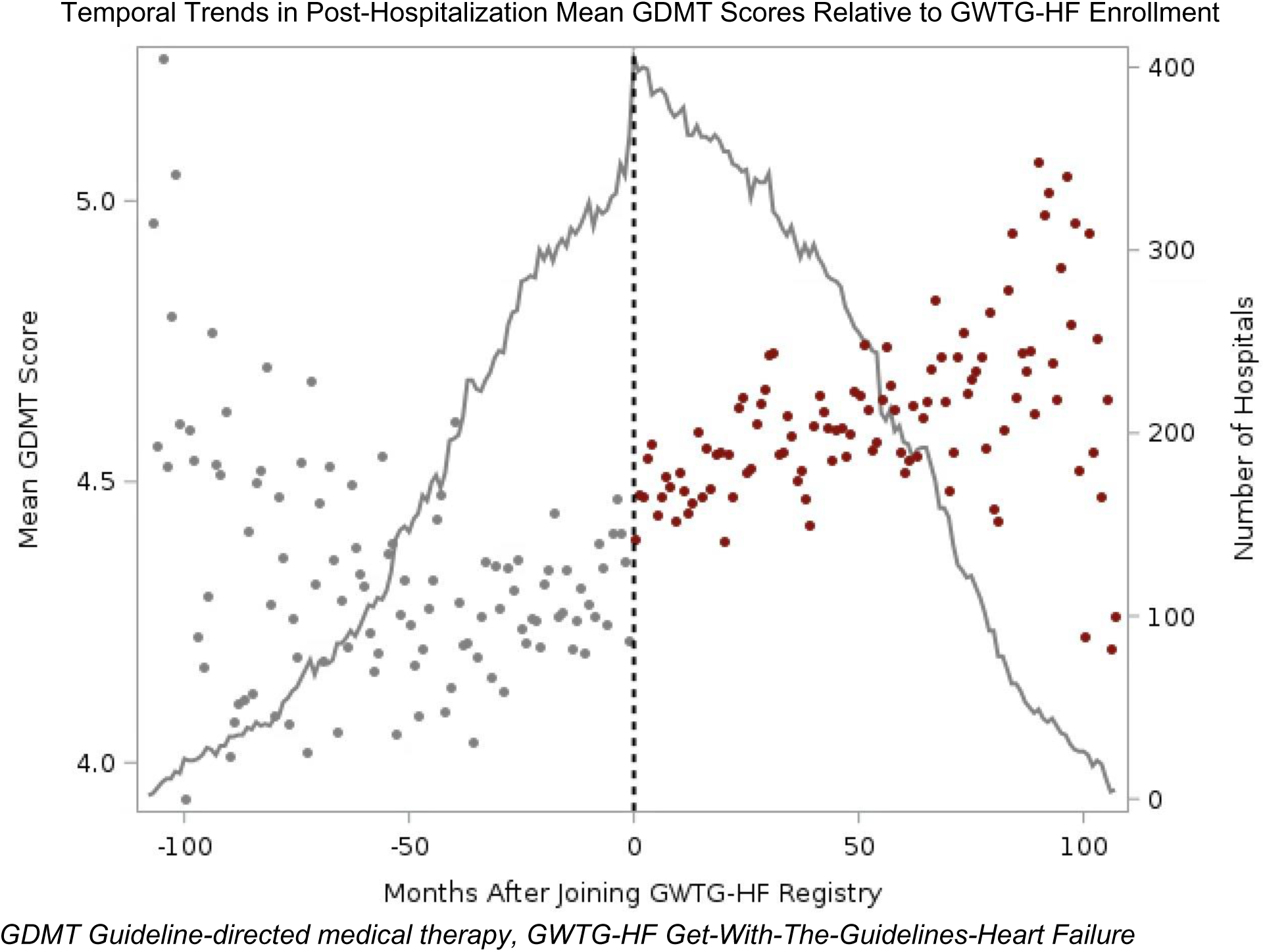
Scatter plot representing the mean post-hospitalization GDMT score across hospitals per month. The vertical dashed line indicates the time of GWTG-HF enrollment (month 0), with observations prior to enrollment (gray) and after enrollment (red). The gray line represents the number of hospitals that contribute data at each time point (right y-axis), reflecting the staggered enrollment of hospitals over the study period.

**Table 3:**
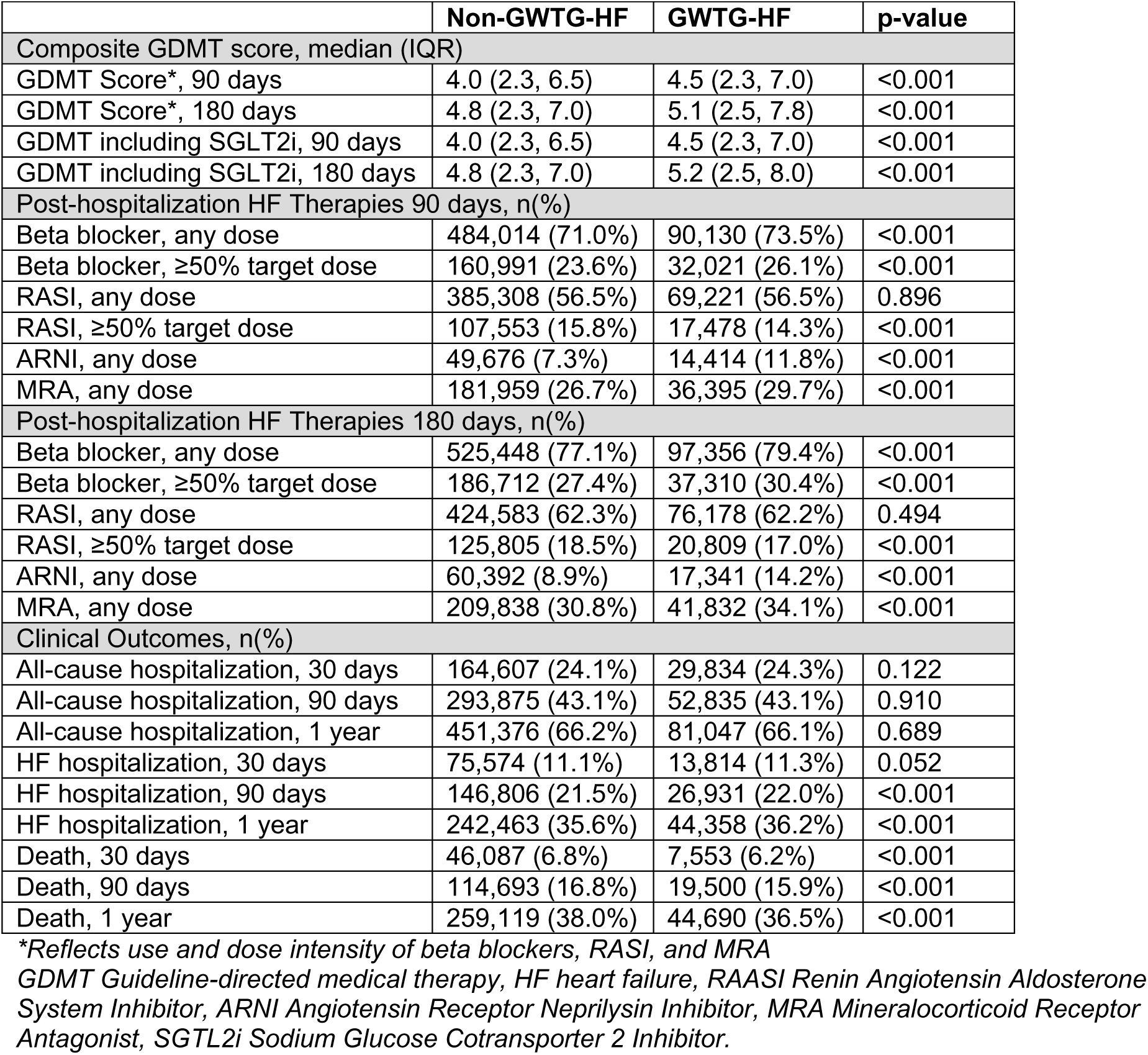
Unadjusted rates of composite GDMT score, individual HF medications, and clinical outcomes stratified by GWTG-HF participation.

|  | Non-GWTG-HF | GWTG-HF | p-value |
| --- | --- | --- | --- |
| Composite GDMT score, median (IQR) |  |  |  |
| GDMT Score*, 90 days | 4.0 (2.3, 6.5) | 4.5 (2.3, 7.0) | <0.001 |
| GDMT Score*, 180 days | 4.8 (2.3, 7.0) | 5.1 (2.5, 7.8) | <0.001 |
| GDMT including SGLT2i, 90 days | 4.0 (2.3, 6.5) | 4.5 (2.3, 7.0) | <0.001 |
| GDMT including SGLT2i, 180 days | 4.8 (2.3, 7.0) | 5.2 (2.5, 8.0) | <0.001 |
| Post-hospitalization HF Therapies 90 days, n(%) |  |  |  |
| Beta blocker, any dose | 484,014 (71.0%) | 90,130 (73.5%) | <0.001 |
| Beta blocker, ≥50% target dose | 160,991 (23.6%) | 32,021 (26.1%) | <0.001 |
| RASI, any dose | 385,308 (56.5%) | 69,221 (56.5%) | 0.896 |
| RASI, ≥50% target dose | 107,553 (15.8%) | 17,478 (14.3%) | <0.001 |
| ARNI, any dose | 49,676 (7.3%) | 14,414 (11.8%) | <0.001 |
| MRA, any dose | 181,959 (26.7%) | 36,395 (29.7%) | <0.001 |
| Post-hospitalization HF Therapies 180 days, n(%) |  |  |  |
| Beta blocker, any dose | 525,448 (77.1%) | 97,356 (79.4%) | <0.001 |
| Beta blocker, ≥50% target dose | 186,712 (27.4%) | 37,310 (30.4%) | <0.001 |
| RASI, any dose | 424,583 (62.3%) | 76,178 (62.2%) | 0.494 |
| RASI, ≥50% target dose | 125,805 (18.5%) | 20,809 (17.0%) | <0.001 |
| ARNI, any dose | 60,392 (8.9%) | 17,341 (14.2%) | <0.001 |
| MRA, any dose | 209,838 (30.8%) | 41,832 (34.1%) | <0.001 |
| Clinical Outcomes, n(%) |  |  |  |
| All-cause hospitalization, 30 days | 164,607 (24.1%) | 29,834 (24.3%) | 0.122 |
| All-cause hospitalization, 90 days | 293,875 (43.1%) | 52,835 (43.1%) | 0.910 |
| All-cause hospitalization, 1 year | 451,376 (66.2%) | 81,047 (66.1%) | 0.689 |
| HF hospitalization, 30 days | 75,574 (11.1%) | 13,814 (11.3%) | 0.052 |
| HF hospitalization, 90 days | 146,806 (21.5%) | 26,931 (22.0%) | <0.001 |
| HF hospitalization, 1 year | 242,463 (35.6%) | 44,358 (36.2%) | <0.001 |
| Death, 30 days | 46,087 (6.8%) | 7,553 (6.2%) | <0.001 |
| Death, 90 days | 114,693 (16.8%) | 19,500 (15.9%) | <0.001 |
| Death, 1 year | 259,119 (38.0%) | 44,690 (36.5%) | <0.001 |

\*Reflects use and dose intensity of beta blockers, RASI, and MRA
GDMT Guideline-directed medical therapy, HF heart failure, RAASI Renin Angiotensin Aldosterone System Inhibitor, ARNI Angiotensin Receptor Neprilysin Inhibitor, MRA Mineralocorticoid Receptor Antagonist, SGLT2i Sodium Glucose Cotransporter 2 Inhibitor.

We evaluated the parallel trends assumption by comparing pre-enrollment temporal trends in GDMT score between hospitals that eventually joined the GWTG-HF registry and those that never joined during the study period. We tested whether the interaction between time and future GWTG-HF participation was associated with GDMT score in the pre-enrollment period. We found no significant differential pre-enrollment trend between groups (Table S1), supporting the parallel trends assumption underlying our primary analysis.

In the primary adjusted analysis, hospitalization at a GWTG-HF hospital was associated with a significantly higher 90-day GDMT score compared with hospitalization at a non-GWTG-HF hospital (+0.15 points; 95% CI 0.12-0.18; p < 0.001) at 90 days. The difference was similar at 180 days (+0.15 point; 95% CI 0.12-0.18, p <0.001). With the alternate slope model, we found each quarter of GWTG-HF participation was associated with a +0.01 increase in post-discharge GDMT score (Table S2).

In the sensitivity analysis for non-parallel baseline trends prior to GWTG-HF hospital participation, the level difference was attenuated to +0.08 (95% CI 0.03-0.12; p<0.001) and +0.06 (95% CI 0.02-0.10, p=0.005) at 90 and 180 days, respectively (Table S2). In another sensitivity analysis, in which we adjusted for hospital characteristics rather than the baseline hospital outcome, we found similar differences in GDMT score between GWTG-HF and non-GWTG-HF hospitals (+0.13 points; 95% CI 0.11-0.15, p<0.001). Lastly, when excluding patients who died prior to the 90-day follow-up, the difference in GDMT score between GWTG-HF and non-GWTG-HF hospitals was +0.16 points (95% CI: 0.13-0.19, p<0.001) at 90 days.

After including SGLT2i in the GDMT score, the median post-discharge score at 90 days was 4.0 [IQR 2.3-6.5] in non-GWTG hospitals and 4.5 [IQR 2.3-7.0] in GWTG hospitals (p<0.001). After adjustment, hospitalization at a GWTG-HF hospital was associated with a significantly higher GDMT score at 90 days post-discharge compared with hospitalization at a non-GWTG-HF hospital (+0.15 points; 95% CI: 0.12-0.18, p < 0.001).

Patients treated at GWTG-HF hospitals were more likely to have filled beta blockers [OR 1.07 (95% CI 1.06-1.09)], RASI [OR 1.02 (95% CI 1.00-1.03)], and MRA [OR 1.13 (95% CI 1.11-1.15)] at 90 days after hospitalization compared with those in non-GWTG-HF hospitals. There was no significant difference in ARNI fills [OR 1.02 (95% CI 0.99-1.04)]. Patients in GWTG-HF hospitals were more likely to achieve ≥50% of target beta blocker doses [OR 1.08 (95% CI 1.06-1.10)], but were less likely to achieve ≥50% of target RASI therapy [OR 0.96 (95% CI 0.93-0.98)] at 90 days (Figure 4).

**Figure 4:**
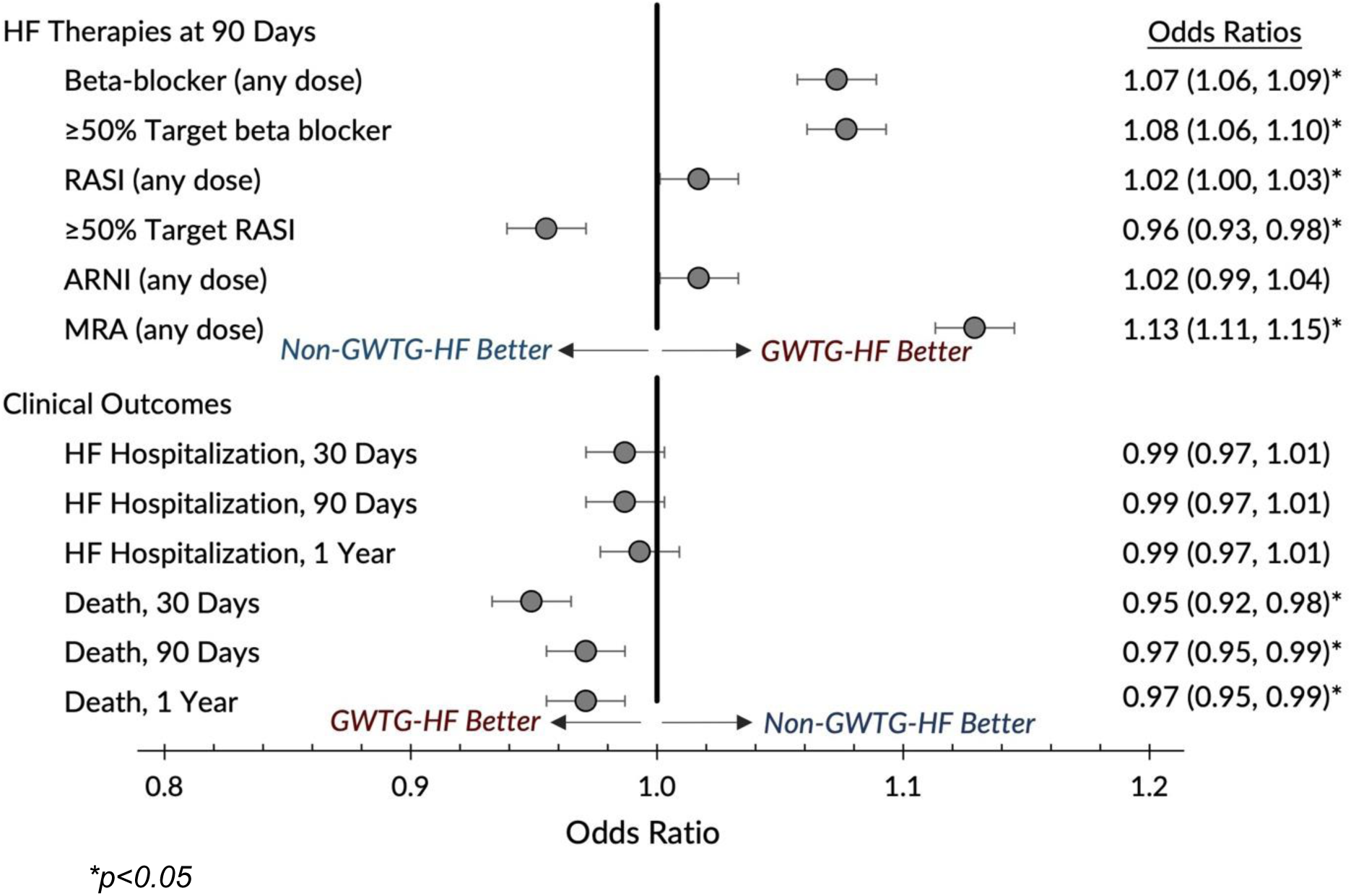
Adjusted odds ratios of HF therapy use at 90 days post-hospitalization and clinical outcomes among patients treated at GWTG-HF versus Non-GWTG-HF hospitals.

In sensitivity analyses accounting for non-parallel trends prior to GWTG-HF participation, these associations were attenuated. The likelihood of MRA fills remained higher in GWTG-HF hospitals [OR 1.07 (95% CI 1.04-1.10)], while the likelihood of RASI decreased slightly [OR 0.97 (95% CI 0.95-1.00)]. There was a small increase in achieving ≥50% target beta blocker therapy [OR 1.03 (95% CI 1.00-1.06)], whereas other differences were no longer statistically significant (Table S3).

#### Clinical Outcomes

At 30 days post-discharge, HF readmission was 11.3% for patients hospitalized at GWTG-HF hospitals and 11.1% for patients hospitalized at non-GWTG-HF (p=0.05). The proportion of all-cause and HF readmissions at 90 days and 1 year was also similar (Table 3). After adjustment, GWTG-HF participation was not associated with differences in HF readmissions at 30 days (OR 0.99; 95% CI 0.97-1.01). We found similar results at 90 days and 1 year. (Figure 4).

At 30 days post-discharge, all-cause mortality was 6.2% for patients hospitalized at GWTG-HF hospitals and 6.8% for patients hospitalized at non-GWTG-HF (p<0.001). At 90 days, mortality was 15.9% vs. 16.8% (p<0.001) and at 1 year, 36.5% vs. 38.0% (p<0.001). After adjustment, GWTG-HF participation was associated with significantly lower post-discharge mortality at 30 days (OR 0.95; 95% CI: 0.92-0.98, 90 days (OR: 0.97; 95% CI: 0.95-0.99), and 1 year (0.97; 95% CI: 0.95-.0.99), all *p*<0.05) (Figure 4). In the sensitivity analysis assuming different trends prior to GWTG-HF participation, this difference was attenuated (Table S4). In an additional model adjusting for 90-day discharge GDMT, the mortality association was no longer significant (Table S5).

## Discussion

Among Medicare beneficiaries hospitalized with HF, we found that hospital participation in the GWTG-HF program was associated with significantly greater use of GDMT among patients with HFrEF. We observed higher odds of post-discharge treatment with beta-blockers, MRA, and RASI following hospital enrollment in the GWTG-HF registry compared with hospitals that did not join the registry. Furthermore, there was a modest but statistically significant associated reduction in all-cause death among patients admitted to GWTG-HF hospitals versus not. Prior studies have shown improvements in process of care metrics at GWTG-HF hospitals, including use of ACE/ARBs for patients with a reduced left ventricular ejection fraction.^4,13^ This study evaluates within-hospital performance changes following GWTG-HF registry participation. This longitudinal perspective helps isolate the association of GWTH-HF participation with quality improvement and provides a contemporary evaluation of the program’s role in promoting GDMT for HF.

Patient demographics, comorbidities, and baseline therapy were overall similar between patients admitted to hospitals that joined the GWTG-HF registry and those that did not. This is an important observation that supports the value of the GWTG-HF registry in analyzing temporal trends in acute HF care in the United States. Nevertheless, there are certain differences in patient and hospital characteristics. Non-GWTG-HF hospitals served twice as many rural patients, a population with higher HF mortality risk and greater barriers to accessing care that could potentially limit HF care and outcomes.^14^ GWTG-HF hospitals also tended to be larger (>400 beds), suggesting possible structural advantages that may have facilitated participation in the program. Targeting recruitment of GWTG-HF to smaller, rural hospitals is an important opportunity to improve quality in populations that already face significant barriers to optimal HF treatment. The AHA has aimed to address this difference via the Rural Health Care Outcomes Accelerator, which aims to increase participation of rural hospitals at no cost in the GWTG registries.^15^

Additionally, GWTG-HF hospitals more frequently had an Accreditation Council for Graduate Medical Education training program and higher CMS quality scores; however, CMS quality ratings were missing for a notable proportion of non-GWTG-HF hospitals (27.5%). Our analysis accounted for differences in hospital characteristics by adjusting for baseline hospital performance and capturing differences in performance following enrollment in the GWTG-HF registry. We also performed a sensitivity analysis adjusting for these characteristics that demonstrated similar findings.

Analyses of individual therapies revealed that GWTG-HF participation was associated with higher use of beta blockers (including achieving ≥50% of target dose), RASI, and MRA. No significant difference was observed in ARNI uptake, likely because it was not the preferred agent in HF guidelines until the later few years of the study period, as well as the impact of its cost to therapy adoption.^16^ Additionally, as an inpatient registry, GWTG-HF may have greater influence on therapy initiation than subsequent uptitration or transition to ARNI therapy. Prior analyses have demonstrated that therapy uptitration is infrequent among patients hospitalized with HF.^17^

While GWTG-HF participation was not associated with a reduction in HF readmission, we observed a modest but statistically significant reduction in all-cause mortality. These findings may reflect that improved GDMT uptake facilitated by the GWTG-HF program may contribute to better survival outcomes. This is further supported by our exploratory analysis, in which adjustment for post-discharge GDMT score attenuated the observed mortality benefit. These findings align with prior studies showing that higher composite GDMT scores are associated with improved survival, more days spent at home, and lower healthcare costs.^18,19^ There may be other improvements in the quality of HF care delivery through the program that translate to improved outcomes. Alternatively, hospital systems that choose to enroll in GWTG-HF may also be engaging in other quality improvement efforts that affect mortality.

We did not observe significant differences in trends in the GDMT score between GWTG-HF and non-GWTG-HF hospitals prior to enrollment in the registry; therefore, we modeled similar baseline trends before GWTG-HF participation. In a sensitivity analysis allowing for different pre-intervention trends in GDMT uptake, the estimated impact of GWTG-HF was attenuated across the GDMT outcome and clinical outcomes. These findings may suggest that hospitals opting into GWTG-HF were already on a trajectory of quality improvement prior to enrollment. Participation in GWTG-HF may still provide the structure, benchmarking, and reporting mechanisms to support and sustain those efforts in improving HF quality of care.

The differences in GDMT score and individual therapies were modest. Future efforts should identify and prioritize the most effective hospital practices within GWTG-HF to increase the impact of the program, and address barriers in adoption for newer therapies such as ARNIs and SGLT2 inhibitors. Prior data suggest that interdisciplinary GDMT outpatient clinics consistently improve therapy optimization, whereas other quality improvement initiatives yield mixed results.^20^ Additionally, systems and payers can use GWTG-HF participation and GDMT score tracking to drive improvement, such as by supporting enrollment of under-resourced hospitals and pairing inpatient checklists with outpatient titration programs. The AHA’s IMPLEMENT-HF initiative is an example of a quality improvement initiative that prioritized post-hospitalization GDMT optimization. The program achieved significant improvements in 4-pillar GDMT and >10% increases in prescriptions for each class of therapy.^21^ These findings underscore the value of HF quality improvement beyond hospitalization and into the outpatient setting where sustained therapy optimization can have the greatest long-term impact.

There are several limitations to this observational study. GWTG-HF hospital participation is voluntary and may be influenced by institutional characteristics such as greater resource availability, leadership engagement, and a culture of quality improvement that both increase the likelihood of participation and the hospital’s capacity to improve care. Our approach accounted for hospital-level differences by adjusting for each hospital’s baseline; however, it does not account for changes in hospital characteristics over time that may have influenced both enrollment in GWTG-HF and improvement in outcomes. As such, we cannot determine whether the observed improvements were a direct result of GWTG-HF program, or if GWTG-HF participation was a marker of hospitals that delivered higher quality care. Notably, the reduction in mortality associated with GWTG-HF participation was not accompanied by a decrease in HF readmission, which may suggest residual confounding contributing to the observed clinical outcomes or mortality as a competing risk with HF hospitalization. Nevertheless, our primary analysis focused on change in the composite GDMT score compared with pre-admission given this is a process metric that is highly under the influence of a treating hospital rather than an outcome measure that is more subject to changes in patient characteristics over time. Using administrative claims data alone, we were unable to account for barriers to therapy uptake (e.g., medication intolerance, contraindications, patient preferences, or cost) and capture device-based HF therapies. While we excluded patients with advanced CKD to account for limited GDMT use in these patients, other clinical factors were not captured. Lastly, the ejection fraction subtype of HF may have been misclassified due to inconsistent or incomplete coding practices. For these reasons, our GDMT rates may differ overall from those observed in analyses of clinical registries, such as the GWTG-HF registry.

## Conclusion

In this national analysis, patients hospitalized at GWTG-HF participating hospitals were more likely to be treated with GDMT and have lower post-discharge mortality, but similar HF rehospitalization rates compared with those hospitalized at non-participating hospitals. These findings underscore the role of structured quality programs in driving the uptake of evidence-based HFrEF therapies and their potential to improve survival.

## Sponsorship

The Get With The Guidelines–Heart Failure (GWTG-HF) program is provided by the American Heart Association. GWTG-HF is sponsored, in part, by Novartis, Boehringer Ingelheim, Novo Nordisk, Bayer and Bristol Myers Squibb

## Data Availability

The data referred to in the manuscript can be made available by individual request.

---

**The RECORD statement – checklist of items, extended from the STROBE statement, that should be reported in observational studies using routinely collected health data.**

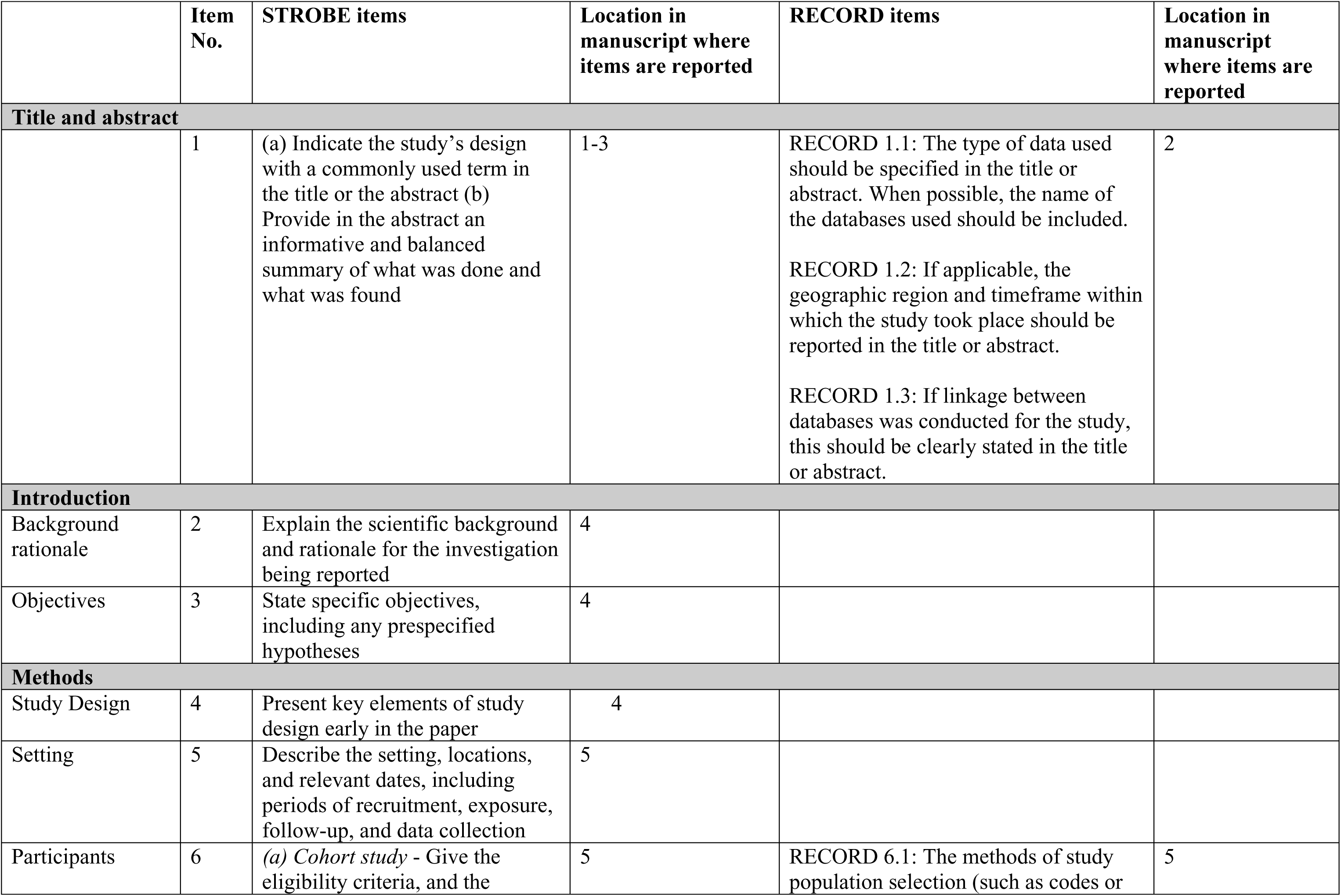
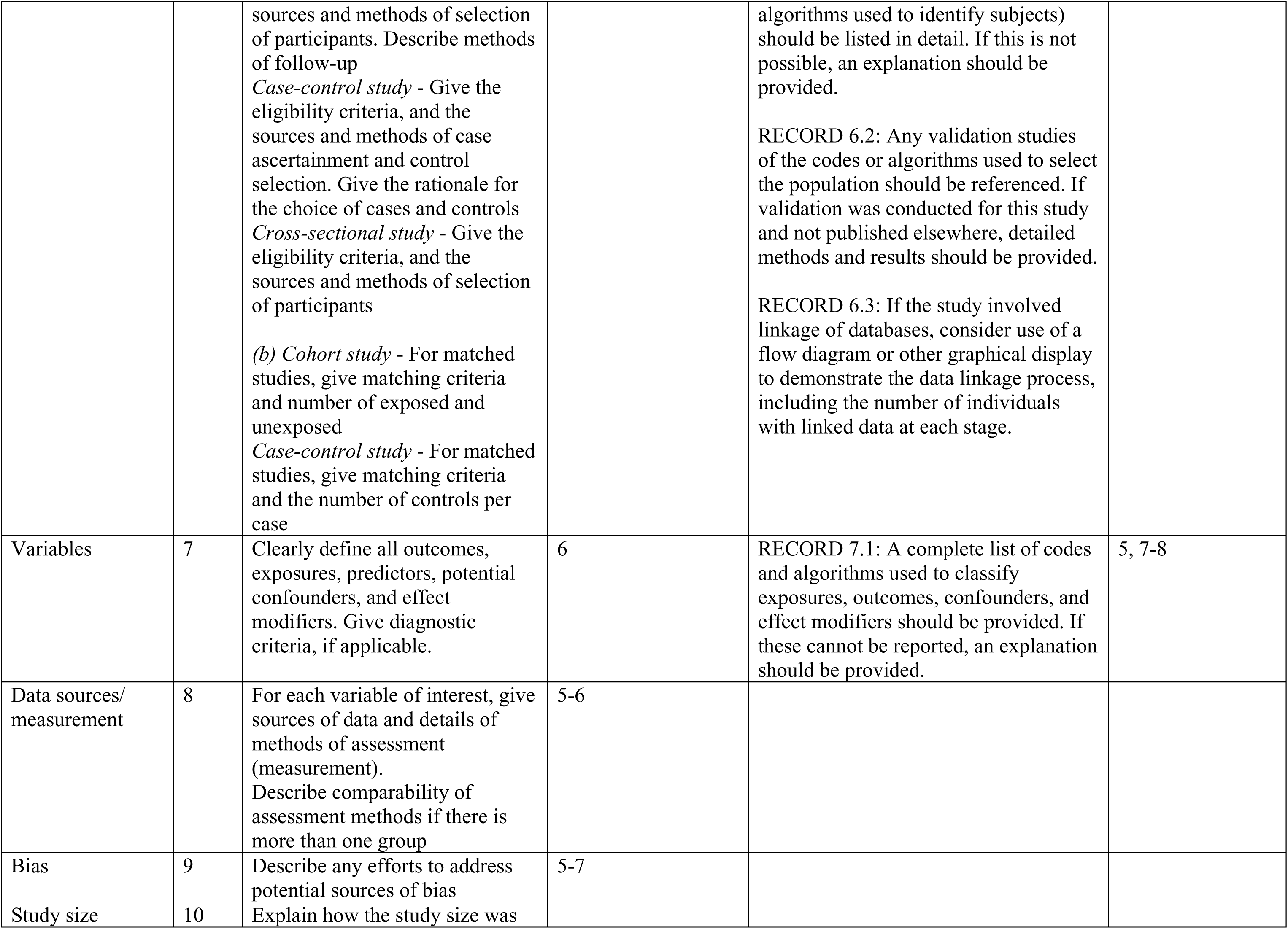
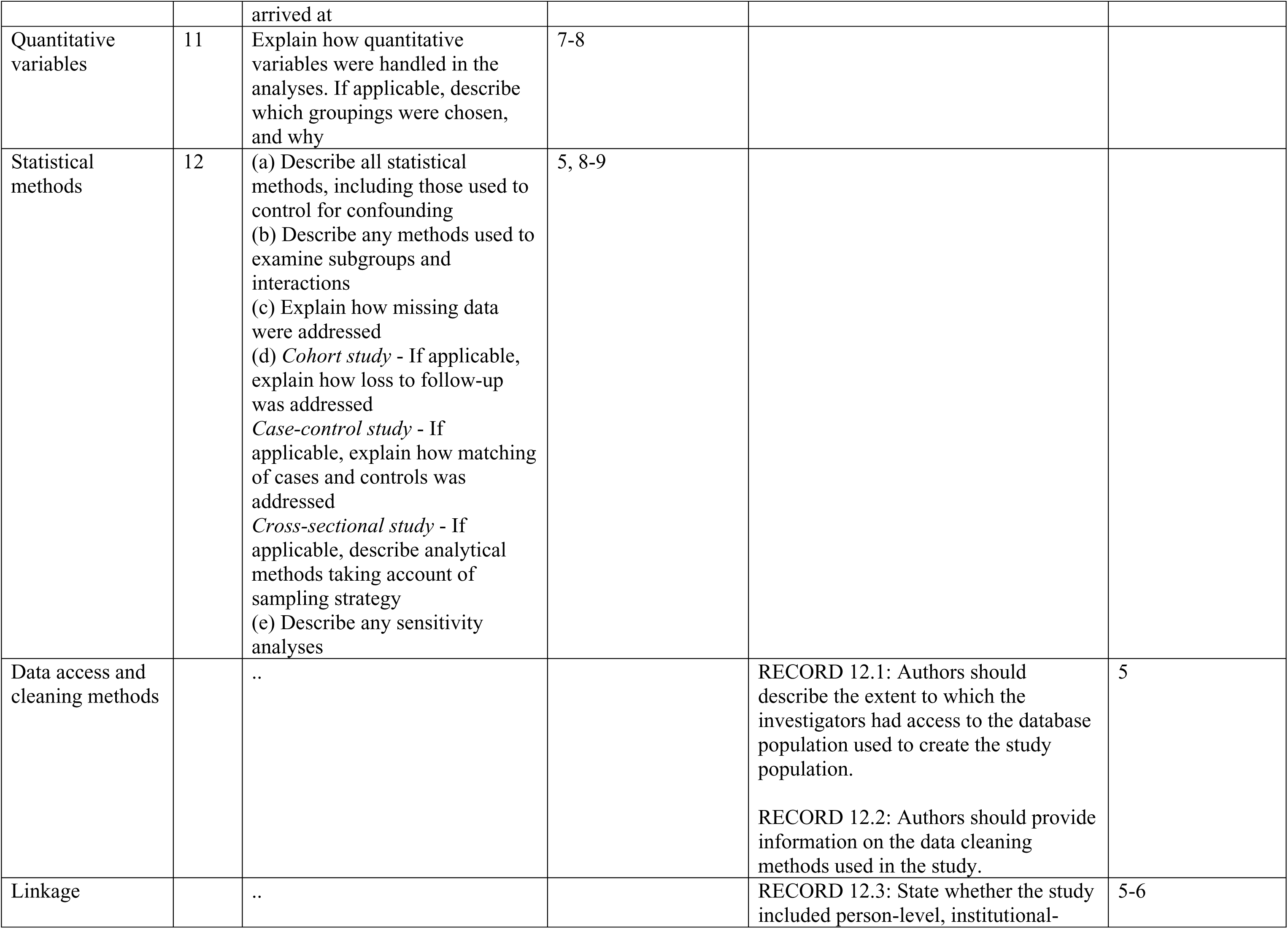
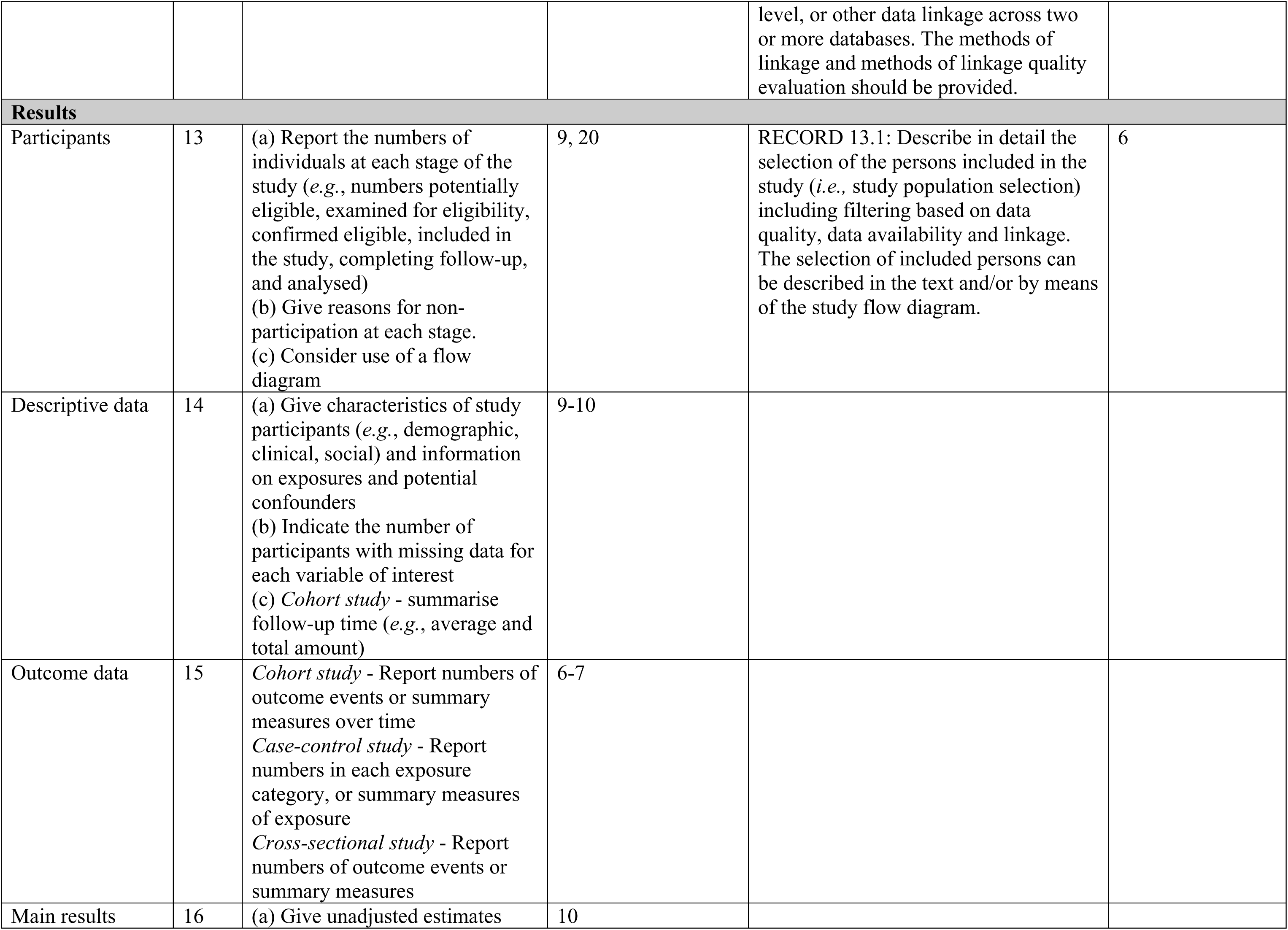
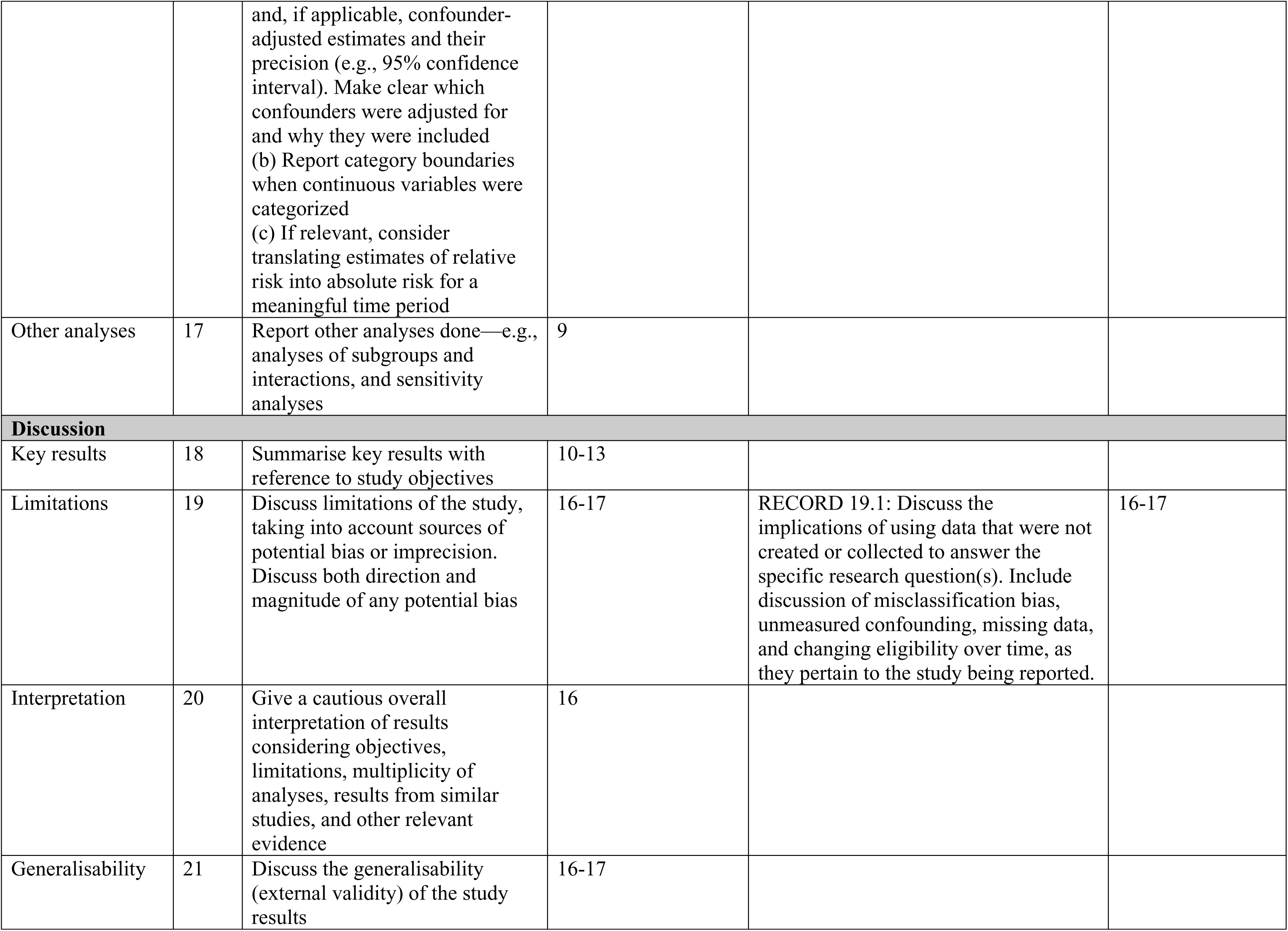
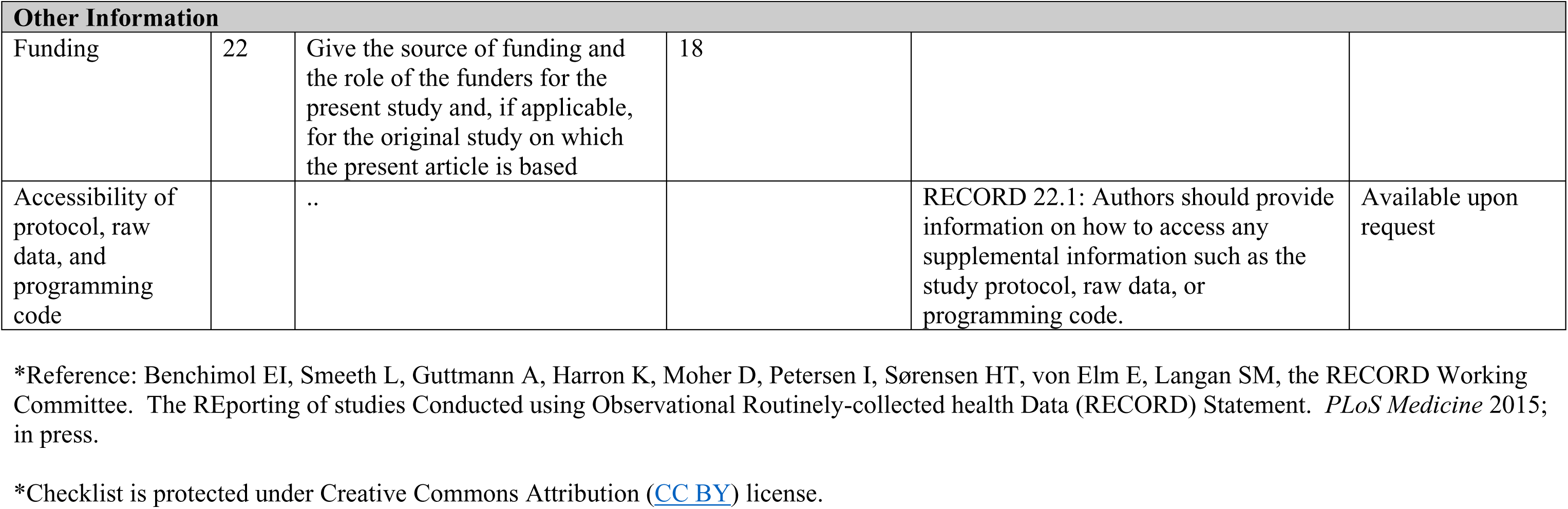

